# Higher burden of cerebral small vessel disease is associated with risk of incident stroke in community dwelling individuals

**DOI:** 10.1101/2024.11.13.24317296

**Authors:** Adlin Pinheiro, Hugo Aparicio, Vasileios Lioutas, Alexa Beiser, Oluchi Ekenze, Charles DeCarli, Sudha Seshadri, Serkalem Demissie, Jose R. Romero

## Abstract

**Background:** Mild manifestations of individual cerebral small vessel disease (CSVD) markers are common and may not denote increased risk, but high CSVD burden identifies individuals at increased risk of stroke and dementia. Scores incorporating multiple individual CSVD markers may better identify a person’s risk. We related a multi-marker CSVD score to risk of incident stroke and compared it with the Framingham Stroke Risk Profile (FSRP) in community-dwelling individuals.

**Methods:** Framingham Heart Study participants aged ≥55 years, free of stroke and dementia and with brain magnetic resonance imaging ratings of CSVD markers were included. A multi-marker CSVD score reflecting increasing CSVD burden was used, assigning one point each for presence of cerebral microbleeds, severe perivascular spaces, extensive white matter hyperintensities, covert brain infarcts, and cortical superficial siderosis. Multivariable Cox proportional hazards regression analyses were used to relate CSVD score to incident stroke.

**Results:** Among 1,154 participants (46% male, mean age 70.9±8.7), 92 (8%) developed stroke over a median follow-up of 8.6 years (Q1-Q3: 5.1-12.5). In models adjusting for age, sex, time interval between clinic exam and MRI, FHS cohort, and FSRP, those with three or more markers had increased risk of stroke (HR: 2.62; 95% CI: 1.17-5.88). In comparison, a 5-percent increase in FSRP was also associated with increased risk (aHR: 1.16; 95% CI: 1.04-1.29). The FSRP and CSVD score had similar model discrimination metrics.

**Interpretation:** Higher CSVD burden is associated with increased risk of stroke, beyond the effect explained by risk factors in the FSRP. These findings support consideration of CSVD burden to identify risk of stroke in community-dwelling individuals for early implementation of preventive strategies.

## INTRODUCTION

Stroke incidence remains high in the U.S. with 795,000 annual cases^1^ and is much higher in developing countries around the world.^2^ Cerebral small vessel disease (CSVD) is recognized as one of the major contributors to stroke and its adverse consequences. Brain magnetic resonance imaging (MRI) has advanced the study of CSVD in subclinical stages, allowing the identification of disease years to decades before stroke events occur, thus offering an opportunity to evaluate individuals at increased risk. Among CSVD markers readily identifiable on MRI are white matter hyperintensities (WMH), covert brain infarcts (CBI), cerebral microbleeds (CMBs), visible (enlarged) perivascular spaces (PVS) and cortical superficial siderosis (cSS).

Although these markers frequently coexist, their presence is heterogenous with prominent interindividual differences. Recently, research efforts have led to the study of “total” CSVD scores which aim to incorporate the presence of multiple markers in an individual to better assess their global contribution to cerebrovascular disease and events. However, most studies have been conducted in clinical samples^3–10^ or small samples,^11–13^ with paucity of data in large, long-term studies of community-dwelling individuals. Such data are needed to further improve public health interventions aiming to ameliorate the consequences of CSVD, including clinical stroke events.

Thus, we studied the relation of a multi-marker CSVD score incorporating all available CSVD measures in relation to incident stroke in the population-based Framingham Heart Study. We also compared its performance for stroke risk prediction against clinical risk factors alone summarized by the Framingham Stroke Risk Profile (FSRP). Our hypothesis was that higher multi-marker CSVD scores would be related to increased stroke risk and capture an individual’s risk as well or better than clinical risk factors alone, which may help identify individuals for targeted preventive therapies.

## METHODS

### Sample

The Framingham Heart Study (FHS) is the longest running prospective community-based cohort study in the United States, now in its 76^th^ year. Participants from multiple generations were enrolled beginning with the Original cohort (N = 5,209) since 1948, followed by the Offspring cohort (N = 5,124) since 1971. Participants have been followed prospectively every two to four years, with 32 examinations in the Original cohort and 10 examinations in the Offspring cohort.

Original and Offspring participants were eligible for the present study if they had at least one available brain MRI with ratings of CSVD markers, had follow up data for incident stroke, and were over the age of 55 at the time of the MRI. If multiple MRIs were available, the earliest MRI was selected. Participants with prevalent stroke, dementia, missing exposure or outcome data, or neurological conditions affecting MRI measurements (e.g. tumor, multiple sclerosis, and head trauma) were excluded. A total of 1,154 participants were included in the final sample (Figure 1).

**Figure 1:**
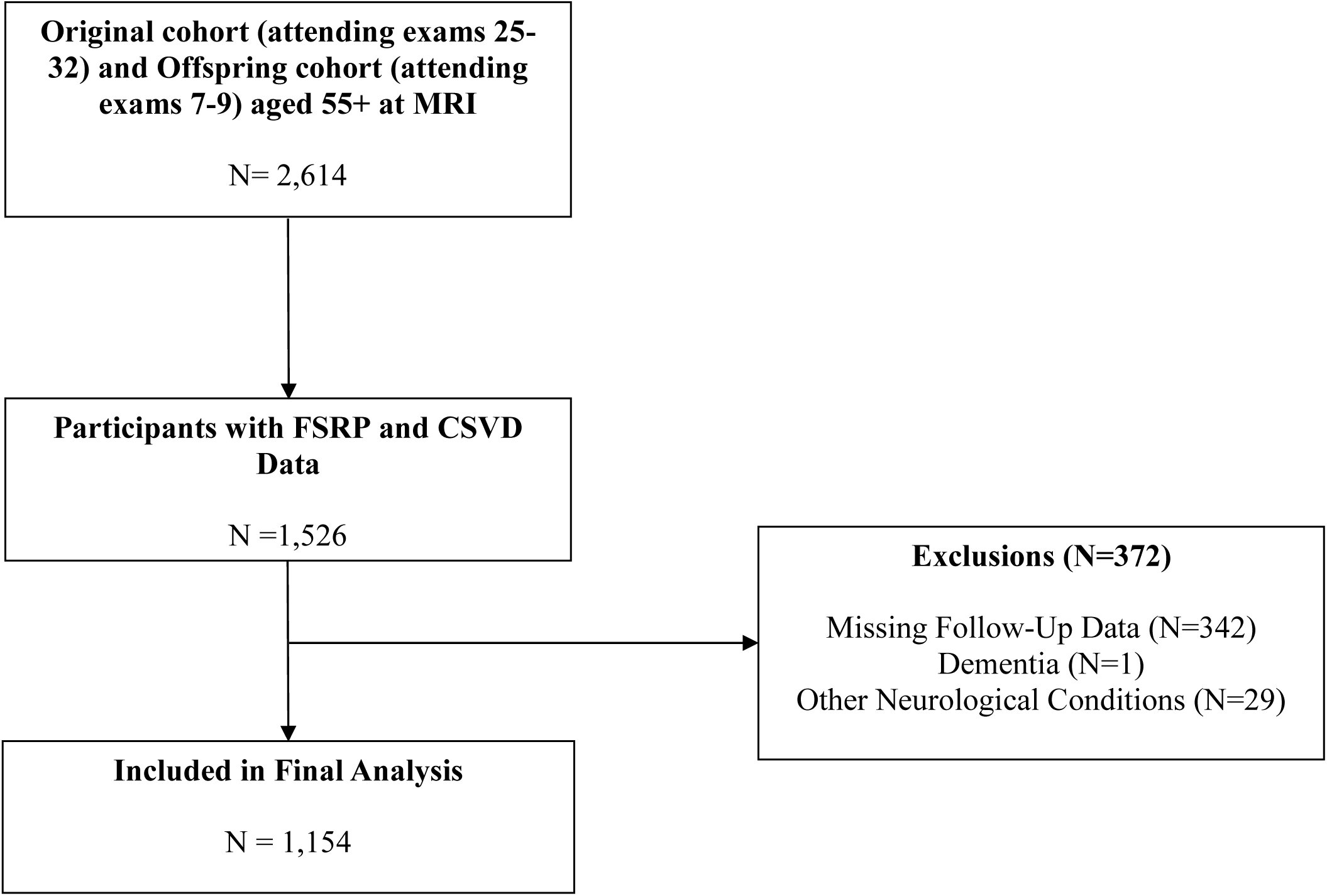
Sample selection flow chart

### Standard Protocol Approvals, Registrations, and Patient Consents

The Institutional Review Board of Boston University Medical Center approved the study protocol, and informed consent was obtained from all subjects.

### Exposures

*Brain MRI.* Brain MRI acquisition measures and image processing methods have been described in detail ^9,10^. Brain MRI have been acquired using a 1T (1999–2005), 1.5T (after 2005) or 3T (after 2010) Magnetom scanner (Siemens Medical, Erlangen, Germany). All ratings and analyses were done blind to the subject’s demographic and clinical characteristics and outcome ascertainment. All CSVD markers were defined and assessed following the Standards for Reporting Vascular Changes on Neuroimaging (STRIVE-2) criteria and published guidelines for individual markers.^14,15^

*Cerebral small vessel disease markers.* Covert brain infarcts were determined based on the size (≥3 mm), location, hyperintensity on T2-weighted images, and imaging characteristics of the lesion (e.g. cerebrospinal fluid signal intensity on subtraction images) using previously described methods.^16^

Cerebral microbleeds were defined and rated using standard criteria^14^ as round or ovoid hypointense lesions on blood sensitive sequences with a diameter up to 10 mm and surrounded by brain parenchyma over at least half the circumference of the lesion. CMB mimics were excluded.^15^

White matter hyperintensities were measured with previously published quantitative methods using T2-weighted and fluid-attenuated inversion recovery (FLAIR) MRI sequences to estimate the volume of WMH.^11^ Presence of extensive WMH was defined as having one standard deviation above an age-grouped mean for WMH volume as a percentage of total brain volume.

MRI-visible perivascular spaces were defined as round, ovoid or linear following the course of penetrating cerebral vessels, with signal intensity resembling cerebrospinal fluid on all sequences, and having a diameter smaller than 3 mm. PVS were assessed in the centrum semiovale (CSO) and basal ganglia (BG) and categorized based on counts: Grade I (1–10), Grade II (11–20), Grade III (20–40), and Grade IV (>40). Because of low prevalence of grade IV cases in FHS participants, we further classified PVS as low burden (grades I and II) or high burden (grades III and IV).

Cortical superficial siderosis was classified as either focal (≤ 3 sulci) or disseminated (≥ 4 sulci) per previously described methods.^17,18^ Intra and inter-rater reliability for all CSVD markers above have been previously reported and considered to range from good to excellent.^16,19,20^

*Multi-marker CSVD score.* We calculated the multi-marker CSVD score using a practical ordinal scale designed for clinical application. The score ranges from 0 to 5, with one point assigned for the presence of each of the following markers: covert brain infarcts, cerebral microbleeds, extensive white matter hyperintensities, high burden of perivascular spaces, and cortical superficial siderosis.

### Outcome

*Assessment of Incident Stroke.* Ongoing surveillance for incident stroke cases includes systematic daily screening of participants admitted to the local hospital in Framingham or other area hospitals, regular FHS examinations, annual heath status updates, ongoing review of Medicare claims data, or report by participants, their family members or primary care physicians via a 24/7 stroke pager (1-800-FHSTROK). All medical records for any emergency room visit, hospitalization or death are reviewed to supplement rigorous screening.

Stroke case adjudication is carried out by a panel of two vascular neurologists evaluating all data, including medical records, imaging studies (reports and images), and diagnostic testing performed to evaluate stroke etiology. Additional data include acute stroke treatment (thrombolytic, mechanical thrombectomy), clinical angiography (CTA, MRA, conventional angiography), evaluation for cardio-embolism (electrocardiogram, trans-thoracic or trans- esophageal echocardiography, cardiac event monitor, implantable loop recorder), carotid ultrasound, surgical procedures, and autopsy. Head CT/MRI is available in more than 90% of cases.

The follow up period extended from the date of the MRI to December 31, 2021.

Censoring for participants who did not experience an incident stroke occurred at the last date known to be stroke free, end of study period, or death from other causes.

*Covariates*. Vascular risk factors were assessed at the closest examination cycle to brain MRI, occurring within 5 years before to one year after the scan. Systolic (SBP) and diastolic (DBP) blood pressures were each taken as the average of the Framingham clinic physician’s two measurements. Hypertension was defined as SBP ≥140mm Hg and/or DBP ≥90mm Hg, or use of antihypertensive medications. Current cigarette smoking was assessed by self-report and defined as any smoking in the year prior to the examination. Diabetes was defined as a random blood glucose ≥200 mg/dl (≥11.1 mmol/L) for the Original cohort, fasting glucose ≥126mg/dl (≥7 mmol/L) for the other cohorts, or use of insulin or oral hypoglycemic medications. Prevalent cardiovascular disease (CVD) included coronary heart disease, heart failure and peripheral arterial disease. Medication use was assessed by self-report and included antiplatelet agents, anticoagulant therapies, and statin use.

*Framingham Stroke Risk Profile (FSRP).* The revised FSRP is a widely used score to predict cardiovascular risk and has been validated in various samples.^21^ The sex-specific score includes the following vascular risk factors to describe a 10-year probability of incident stroke: age, systolic blood pressure, use of antihypertensive medications, prevalent cardiovascular disease, current smoking status, current or previous atrial fibrillation, and diabetes mellitus (DM).

### Statistical Analysis

Descriptive characteristics for baseline demographic, clinical and imaging variables included frequencies and percentages for categorical variables and mean and standard deviation for continuous variables. Due to few participants having more than three CSVD markers, the multi-marker CSVD score was re-categorized into the following: 0 (no CSVD biomarker, reference group), 1 (one biomarker), 2 (two biomarkers) and 3+ (three or more biomarkers) for multivariable analyses. We also evaluated a second score collapsing two or more markers into one group to determine whether results depend on choice of cut point.

Incidence rates (per 1,000 person-years) were calculated for incident stroke by dividing the total number of new stroke events by the total follow-up time and multiplying by 1,000.

Three multivariable Cox regression analyses were then used to relate the multi-marker CSVD score to incident stroke: model 1 adjusted for age, sex, FHS cohort, time interval between MRI and clinic exam; model 2 additionally adjusted for smoking, systolic blood pressure, prevalent CVD, diabetes, atrial fibrillation, and medication use; model 3 adjusted for the covariates in model 1 and FSRP. Effect modification by age and sex was also evaluated in stratified analyses. Lastly, we related FSRP to incident stroke adjusting for the covariates in model 1. Hazard ratios for the FSRP were expressed as a 5% increase in the 10-year probability of stroke.

We compared models with the multi-marker CSVD score and the FSRP using Harrell’s c-statistic, a measure analogous to the area under the curve of a receiver-operating curve (ROC), allowing assessment of discrimination of participants with longer versus shorter event-free survival.^22^ Statistical analyses were performed using SAS version 9.4 (Cary, NC), and a p < 0.05 was considered statistically significant.

Data from this study may be shared with qualified investigators following FHS procedures outlined at https://www.framinghamheartstudy.org/.

## RESULTS

*Sample characteristics.* Participants in the sample included were middle-aged adults with similar proportion of men and women (46% and 54% respectively). The most prevalent vascular risk factor was hypertension. The presence of vascular risk factors increased as the multi-marker CSVD score increased (Table 1).

**Table 1.** Sample Characteristics.

| Clinical Characteristics | Overall<br>N=1,154 | CSVD Score |  |  |  |  |
| --- | --- | --- | --- | --- | --- | --- |
|  |  | 0<br>(N=590) | 1<br>(N=352) | 2<br>(N=164) | 3<br>(N=42) | 4<br>(N=6) |
| Age at MRI, years, mean (SD) | 70.9 (8.7) | 68.2 (7.9) | 73.1 (8.4) | 74.1 (8.7) | 76.6 (8.1) | 80.6 (7.1) |
| Time between clinic exam and MRI, years, median (Q1, Q3) | 0.9 (0.1, 1.7) | 1.0 (0.1, 1.8) | 0.8 (0.1, 1.7) | 0.7 (0.1, 1.6) | 0.7 (0.0, 1.3) | 0.4 (-0.8, 1.1) |
| Men, n (%) | 527 (46) | 276 (47) | 155 (44) | 75 (46) | 20 (48) | 1 (17) |
| Cohort, n (%) |  |  |  |  |  |  |
| Original | 77 (7) | 15 (3) | 35 (10) | 18 (11) | 7 (17) | 2 (33) |
| Offspring | 1077 (93) | 575 (97) | 317 (90) | 146 (89) | 35 (83) | 4 (67) |
| Follow up period, years, median (Q1, Q3) | 8.6 (5.1, 12.5) | 8.8 (5.3, 12.6) | 7.5 (5.0, 12.2) | 8.3 (4.8, 12.2) | 5.7 (3.8, 11.2) | 8.5 (4.6, 13.1) |
| Education, n (%) |  |  |  |  |  |  |
| Less than high school | 45 (4) | 16 (3) | 16 (5) | 9 (5) | 4 (10) | 0 (0) |
| High school | 317 (27) | 151 (26) | 94 (27) | 59 (36) | 8 (19) | 5 (83) |
| Some college | 351 (30) | 179 (30) | 117 (33) | 42 (26) | 12 (29) | 1 (17) |
| College | 439 (38) | 244 (41) | 124 (35) | 54 (33) | 17 (40) | 0 (0) |
| APOE4+, n (%) | 262 (23) | 126 (21) | 74 (21) | 48 (29) | 12 (29) | 2 (33) |
| **Vascular risk factors** |  |  |  |  |  |  |
| Systolic blood pressure, mmHg, mean (SD) | 129.0 (17.6) | 126.2 (16.1) | 130.1 (18.1) | 134.2 (18.3) | 135.6 (20.7) | 151.2 (18.9) |
| Diastolic Blood pressure, mmHg, mean (SD) | 72.3 (10) | 73.4 (9.4) | 70.6 (9.7) | 71.9 (11.5) | 71.1 (11.5) | 80.8 (11.5) |
| Hypertension<sup>a</sup>, n (%) | 675 (58) | 293 (50) | 232 (66) | 115 (70) | 30 (71) | 5 (83) |
| Hypertension treatment, n (%) | 566 (49) | 244 (41) | 201 (57) | 91 (55) | 25 (60) | 5 (83) |
| Current smoker, n (%) | 74 (6) | 38 (6) | 27 (8) | 6 (4) | 2 (5) | 1 (17) |
| Diabetes, n (%) | 173 (15) | 83 (14) | 54 (15) | 29 (18) | 4 (10) | 3 (50) |
| Diabetes treatment, n (%) | 107 (9) | 48 (8) | 35 (10) | 19 (12) | 3 (7) | 2 (33) |
| Anti-lipid treatment, n (%) | 507 (44) | 250 (42) | 167 (47) | 67 (41) | 19 (45) | 4 (67) |
| Prevalent cardiovascular disease, n (%) | 192 (17) | 68 (12) | 81 (23) | 29 (18) | 10 (24) | 4 (67) |
| **MRI Markers of CSVD** |  |  |  |  |  |  |
| Severe perivascular spaces, n (%) | 395 (34) | 0 (0) | 202 (57) | 147 (90) | 40 (95) | 6 (100) |
| Extensive white matter hyperintensities, n (%) | 155 (13) | 0 (0) | 38 (11) | 79 (48) | 32 (76) | 6 (100) |
| Cerebral microbleed, n (%) | 124 (11) | 0 (0) | 52 (15) | 44 (27) | 22 (52) | 6 (100) |
| Covert brain infarct, n (%) | 154 (13) | 0 (0) | 60 (17) | 58 (35) | 30 (71) | 6 (100) |
| Cortical superficial siderosis, n (%) | 2 (0) | 0 (0) | 0 (0) | 0 (0) | 2 (5) | 0 (0) |

CSVD = cerebral small vessel disease
\*Values are mean (SD) for continuous variables and n (%) for categorical variables.
<sup>b</sup> Hypertension is defined as SBP ≥ 140 mmHg or DBP ≥ 90 mmHg and/or use of antihypertensive medication

The prevalence of individual CSVD markers and contribution to the multi-marker score is shown in Figure 2. The multi-marker CSVD score prevalence was 30.5% for only one marker, 14.2% for two markers, 3.6% for three markers, and 0.5% for four markers; no participants had all five markers. In participants with one marker of CSVD, the main contributing marker was high burden of PVS followed by CBI, CMB, and extensive WMH. Similarly, PVS was the main contributing marker for CSVD scores two and three or greater.

**Figure 2:**
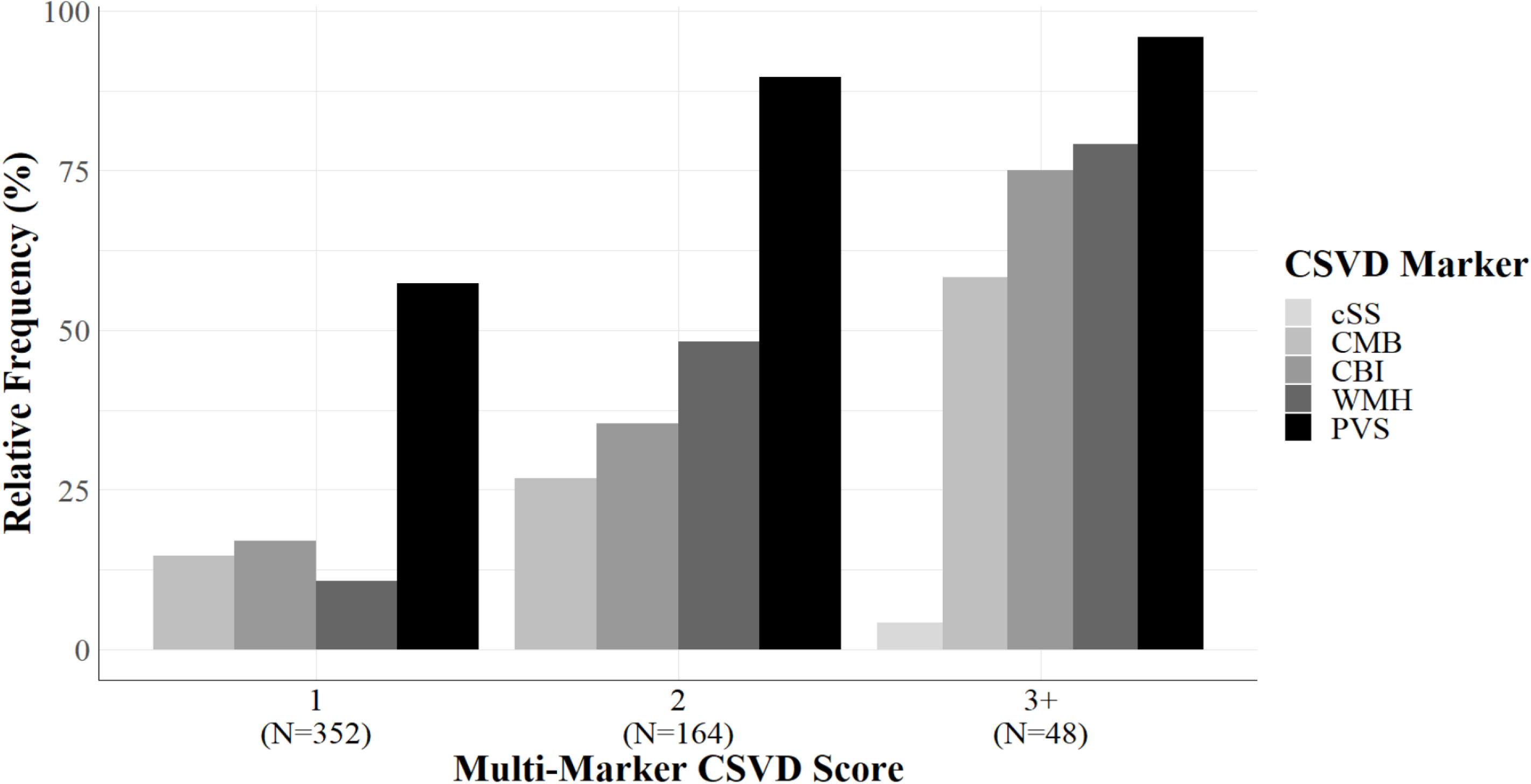
Proportion of CSVD markers comprising total CSVD score

Over a median follow-up period of 8.2 years (Q1-3: 5.1-12.5), there were 92 incident stroke cases (8% over the study period). Crude incidence rates for stroke increased with higher multi-marker CSVD scores (Table 3).

**Table 3.** Incidence stroke rates by multi-marker CSVD score.

| Outcome | Measure | Total | Multi-marker CSVD score |  |  |  |  |
| --- | --- | --- | --- | --- | --- | --- | --- |
|  |  |  | 0<br>(N=590) | 1<br>(N=352) | 2<br>(N=164) | 3<br>(N=42) | 4<br>(N=6) |
| Incident Stroke |  |  |  |  |  |  |  |
|  | N events | 92 | 31 | 35 | 18 | 6 | 2 |
|  | Follow-up time<br>(person-years) | 9968.34 | 5394.52 | 2899.97 | 1379.47 | 291.04 | 48.44 |
|  | Incidence rate<br>(95% CI) | 9.23<br>(7.52, 11.32) | 5.79<br>(4.08, 8.24) | 12.07<br>(8.67, 16.81) | 13.05<br>(8.22, 20.71) | 20.62<br>(9.26, 45.89) | - |

CI = confidence interval; CSVD = cerebral small vessel disease
Incidence rates per 1,000 person-years. Rates are not presented where there were fewer than 5 events.

*Multivariable analyses*. Participants with CSVD score three or greater had significantly higher risk of stroke after adjusting for cardiovascular risk factors (HR: 2.81; 95% CI: 1.24-6.36) and FSRP (HR: 2.62; 95% CI: 1.17-5.88) compared to participants without any CSVD markers. When we used the three-category CSVD score, we similarly observed that participants with two or more markers had significantly higher risk of stroke across all models.

In addition, each 5-unit increase in FSRP was associated with a modest increase in risk (HR: 1.25; 95% CI: 1.17-1.33). However, after adjusting for age, sex, FHS cohort, and time interval, the effect of FSRP was attenuated (HR: 1.16; 95% CI: 1.04-1.29).

*Effect modification.* In stratified analyses evaluating the role of age, we observed higher stroke risk in individuals ≥65 across all multi-marker CSVD scores (Table 4). Although there were no participants younger than 65 years with two or more CSVD markers who developed stroke, those with a multi-marker score of one had significantly higher risk of stroke compared with those without any CSVD markers (HR: 3.21; 95% CI: 1.24, 8.31). Some heterogeneity in stroke risk was seen across multi-marker CSVD scores but overall higher risk was observed in women.

**Table 4:** Cox Proportional Hazard Analysis of Risk of Incident Stroke according to multi-marker CSVD score.

|  |  | **HR (95% CI)** |  |  |
| --- | --- | --- | --- | --- |
| **Three-level CSVD score** | **N** | **Model 1** | **Model 2** | **Model 3** |
| 0 | 590 | Ref | Ref | Ref |
| 1 | 352 | 1.60 (0.98, 2.63) | 1.69 (1.02, 2.82) | 1.56 (0.95, 2.58) |
| 2 or greater | 212 | 1.77 (1.02, 3.05) | 1.93 (1.11, 3.37) | 1.78 (1.03, 3.08) |
| C-statistic |  | 0.69 | 0.70 | 0.70 |
| **Four-level CSVD score** |  |  |  |  |
| 0 | 590 | Ref | Ref | Ref |
| 1 | 352 | 1.61 (0.98, 2.64) | 1.70 (1.02, 2.84) | 1.57 (0.95, 2.58) |
| 2 | 164 | 1.57 (0.86, 2.87) | 1.71 (0.93, 3.14) | 1.57 (0.86, 2.87) |
| 3 or greater | 48 | 2.50 (1.11, 5.61) | 2.81 (1.24, 6.36) | 2.62 (1.17, 5.88) |
| C-statistic |  | 0.69 | 0.71 | 0.70 |
|  |  | **Unadjusted HR (95% CI)** | **Model 1 HR (95% CI)** |  |
| Framingham Stroke Risk Profile* |  | 1.25 (1.17, 1.33) | 1.16 (1.04, 1.29) |  |
| C-statistic |  | 0.66 | 0.68 |  |

The three level CSVD score groups individuals with 2 or more CSVD markers in one category, while the four level one keeps those with 2 markers as a separate group and groups individuals with a score of 3 or greater together.
CI = confidence interval; CSVD = cerebral small vessel disease; HR = hazard ratio
Model 1 covariates: age, sex, FHS cohort, and time interval between MRI and exam cycle
Model 2 covariates: Model 1 covariates, smoking, systolic blood pressure, prevalent CVD, diabetes, atrial fibrillation, and medication use
Model 3 covariates: age, sex, FHS cohort, time interval between MRI and exam cycle, and FSRP score

**Table 5.**
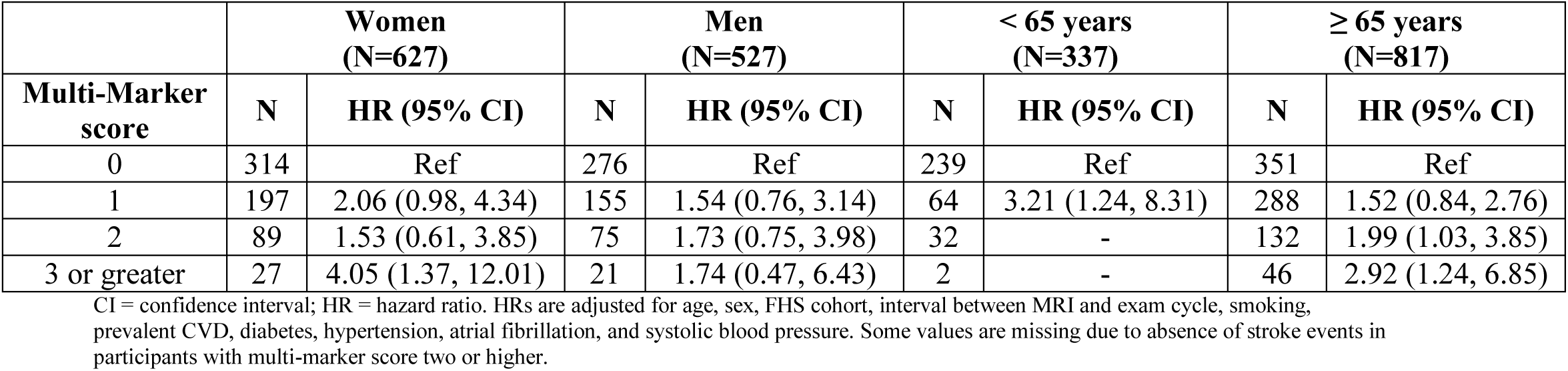
Effect modification by sex and age on the association of multi-marker CSVD score and incident stroke risk.

|  | **Women**<br>**(N=627)** |  | **Men**<br>**(N=527)** |  | **< 65 years**<br>**(N=337)** |  | **≥ 65 years**<br>**(N=817)** |  |
| --- | --- | --- | --- | --- | --- | --- | --- | --- |
| **Multi-Marker**<br>**score** | **N** | **HR (95% CI)** | **N** | **HR (95% CI)** | **N** | **HR (95% CI)** | **N** | **HR (95% CI)** |
| 0 | 314 | Ref | 276 | Ref | 239 | Ref | 351 | Ref |
| 1 | 197 | 2.06 (0.98, 4.34) | 155 | 1.54 (0.76, 3.14) | 64 | 3.21 (1.24, 8.31) | 288 | 1.52 (0.84, 2.76) |
| 2 | 89 | 1.53 (0.61, 3.85) | 75 | 1.73 (0.75, 3.98) | 32 | - | 132 | 1.99 (1.03, 3.85) |
| 3 or greater | 27 | 4.05 (1.37, 12.01) | 21 | 1.74 (0.47, 6.43) | 2 | - | 46 | 2.92 (1.24, 6.85) |

CI = confidence interval; HR = hazard ratio. HRs are adjusted for age, sex, FHS cohort, interval between MRI and exam cycle, smoking, prevalent CVD, diabetes, hypertension, atrial fibrillation, and systolic blood pressure. Some values are missing due to absence of stroke events in participants with multi-marker score two or higher.

*Model comparisons.* Model performance comparing CSVD score, CSVD score adjusted for individual components of the FSRP, and CSVD score adjusted for the FSRP were similar (c=0.69, 0.71, and 0.70, respectively). Additionally, CSVD score and FSRP had similar c- statistics using the covariates in model 1 (c=0.69 and 0.68, respectively).

## Discussion

In this large cohort study of asymptomatic community-dwelling individuals, we observed participants with higher multi-marker CSVD scores were at increased risk of stroke, with greater risk as the number of single CSVD measures increased. The CSVD score signaled increased risk even after adjustment for vascular risk factors as individual covariates or summarized in the FSRP, indicating that CSVD score denotes risk independent of the FSRP.

Both CSVD score and FSRP had similar model performance metrics for stroke risk prediction when comparing c-statistics for both models. This suggests that the CSVD score captures additional or different aspects of stroke risk that are as informative of risk prediction as the FSRP.

There has been an increase in interest in the study of multiple measures of cerebral small vessel disease summarized in simple, practical scores. It has been observed that total CSVD scores are associated with higher risk of recurrent stroke, dementia and mortality.^23,24^ Similarly, in patients with cognitive disorders, total CSVD scores are related to higher risk of progression of cognitive disorders and mortality. Expanding findings from the clinical studies mentioned, our study also showed that asymptomatic individuals in the community are at an increased risk of incident stroke when subclinical CSVD is detected years prior to the event. The risk is independent of vascular risk factor assessments.

Vascular brain injury is a key factor in the development of neurological disorders involving brain disease. The neurovascular unit is a concept that highlights the integration of neuronal, glial and vascular functions for brain health, thus it is not surprising that risk of adverse neurological outcomes is increased when high burden of cerebral small vessel disease is present. Our findings highlight the opportunity to intervene when such findings are detected in subclinical stages. While there is still no specific treatment for CSVD in subclinical stages, its strong association with vascular risk factors, particularly hypertension, calls for implementation of treatments and monitoring of its effectiveness in such patients.

Clinical trials now incorporate subclinical CSVD in their outcomes and it is likely that additional data will become available to guide clinical decisions in the near future. For instance, the Lacunar Intervention Trial-2 (LACI-2)^25^ randomized clinical trial assessed treatments that target endothelial dysfunction in patients with CSVD-related stroke, showing promising results for risk reduction for recurrent stroke and cognitive impairment.

It is important to note that we labeled our scores as multi-marker CSVD scores because the term “total” suggests the inclusion of all possible CSVD markers. While this comprehensive approach is desirable, evolving knowledge and detection methods for CSVD may lead to new measures that must be integrated with traditional ones. Therefore, “total” scores today may differ significantly from “total” scores in the near future.

Our exploratory stratified analyses showed higher risk of stroke in women with high CSVD scores compared to men. In addition, risk of stroke in participants with one CSVD marker was higher in those less than 65 years of age. Comparison of vascular risk factors between men and women showed that women had higher total cholesterol (199.3 vs 173.9 mg/dL), low density lipoprotein – LDL (109.8 vs 99.9mg./dL) and lower use of lipid lowering treatments (39 vs 49%); similar findings were noted in the younger age group where individuals younger than 65 years had higher mean levels of total cholesterol (196 vs 184 mg/dL) and low density lipoprotein – LDL (113.7 vs 101.8) and lower use of lipid lowering therapy (30 vs. 50%); while it remains a speculation it is possible that treatment effects and level of control of some vascular risk factors over time may explain this difference in stroke risk. However, this requires further prospective studies to assess differences in CSVD burden and outcomes in men and women, young and old individuals.

Our study has several strengths including the inclusion of a large sample, blinded assessments of exposure and outcomes, comprehensive longitudinal follow-up for incident stroke with high retention, and accurate and reliable ascertainment of exposures, confounders and outcomes. Limitations include a small number of participants with the highest CSVD scores and the predominantly white racial composition of FHS participants, thus restricting generalization to other racial groups. In addition, selection of participants based on availability of CSVD markers may have resulted in the inclusion of generally healthier participants which may underestimate risk in the included sample.

## CONCLUSION

Multi-marker CSVD scores capture the heterogeneity of CSVD manifestations and signal increased risk of incident stroke when detected in subclinical stages, even in community-dwelling individuals. These scores are independent of the FSRP, capturing different aspects of stroke risk not accounted for by the FSRP, and had similar model discrimination performance to the FSRP. Further studies are needed to assess implementation of preventive measures in patients with high burden of subclinical CSVD.

## Source of Funding

This work (design and conduct of the study, collection and management of the data) was supported by the Framingham Heart Study’s National Heart, Lung, and Blood Institute contract (N01-HC-25195; HHSN268201500001I) and by grants from the National Institute of Neurological Disorders and Stroke (R01-NS017950-37, R21 NS135268), the National Institute on Aging (R01 AG059725; AG008122; AG054076; K23AG038444; R03 AG048180-01A1; AG033193); NIH grant (P30 AG010129).

---

**Supplementary Table 1:**

| Clinical Characteristics | Overall<br>N=1,154 | CSVD Score |  |  |  |
| --- | --- | --- | --- | --- | --- |
|  |  | 0<br>(N=590) | 1<br>(N=352) | 2<br>(N=164) | 3+<br>(N=48) |
| Age at exam closest to MRI, years, mean (SD) | 69.4 (8.8) | 66.6 (8.0) | 71.6 (8.6) | 72.7 (8.7) | 75.8 (8.5) |
| Age at MRI, years, mean (SD) | 70.9 (8.7) | 68.2 (7.9) | 73.1 (8.4) | 74.1 (8.7) | 77.1 (8.1) |
| Time between clinic exam and MRI, years, median (Q1, Q3) | 0.9 (0.1, 1.7) | 1.0 (0.1, 1.8) | 0.8 (0.1, 1.7) | 0.7 (0.1, 1.6) | 0.6 (0.0, 1.3) |
| Men, n (%) | 527 (46) | 276 (47) | 155 (44) | 75 (46) | 21 (44) |
| Cohort, n (%) |  |  |  |  |  |
| Original | 77 (7) | 15 (3) | 35 (10) | 18 (11) | 9 (19) |
| Offspring | 1077 (93) | 575 (97) | 317 (90) | 146 (89) | 39 (81) |
| Follow up period, years, median (Q1, Q3) | 8.6 (5.1, 12.5) | 8.8 (5.3, 12.6) | 7.5 (5.0, 12.2) | 8.3 (4.8, 12.2) | 5.7 (4.0, 11.6) |
| Education, n (%) |  |  |  |  |  |
| Less than high school | 45 (4) | 16 (3) | 16 (5) | 9 (5) | 4 (8) |
| High school | 317 (27) | 151 (26) | 94 (27) | 59 (36) | 13 (27) |
| Some college | 351 (30) | 179 (30) | 117 (33) | 42 (26) | 13 (27) |
| College | 439 (38) | 244 (41) | 124 (35) | 54 (33) | 17 (35) |
| APOE4+, , n (%) | 262 (23) | 126 (21) | 74 (21) | 48 (29) | 14 (29) |
| ***Vascular risk factors*** |  |  |  |  |  |
| Systolic blood pressure, mmHg, mean (SD) | 129.0 (17.6) | 126.2 (16.1) | 130.1 (18.1) | 134.2 (18.3) | 137.6 (21) |
| Diastolic Blood pressure, mmHg, mean (SD) | 72.3 (10.0) | 73.4 (9.4) | 70.6 (9.7) | 71.9 (11.5) | 72.4 (11.8) |
| Hypertension<sup>a</sup>, n (%) | 675 (58) | 293 (50) | 232 (66) | 115 (70) | 35 (73) |
| Hypertension treatment, n (%) | 566 (49) | 244 (41) | 201 (57) | 91 (55) | 30 (63) |
| Current smoker, n (%) | 74 (6) | 38 (6) | 27 (8) | 6 (4) | 3 (6) |
| Diabetes, n (%) | 173 (15) | 83 (14) | 54 (15) | 29 (18) | 7 (15) |
| Diabetes treatment, n (%) | 107 (9) | 48 (8) | 35 (10) | 19 (12) | 5 (10) |
| Anti-lipid treatment, n (%) | 507 (44) | 250 (42) | 167 (47) | 67 (41) | 23 (48) |
| Prevalent cardiovascular disease | 192 (17) | 68 (12) | 81 (23) | 29 (18) | 14 (29) |
| ***MRI Markers of CSVD*** |  |  |  |  |  |
| Severe perivascular spaces, n (%) | 395 (34) | 0 (0) | 202 (57) | 147 (90) | 46 (96) |
| Extensive white matter hyperintensities, n (%) | 155 (13) | 0 (0) | 38 (11) | 79 (48) | 38 (79) |
| Cerebral microbleed, n (%) | 124 (11) | 0 (0) | 52 (15) | 44 (27) | 28 (58) |
| Covert brain infarct, n (%) | 154 (13) | 0 (0) | 60 (17) | 58 (35) | 36 (75) |
| Cortical superficial siderosis, n (%) | 2 (0) | 0 (0) | 0 (0) | 0 (0) | 2 (4) |

## Notes

### Competing Interest Statement

The authors have declared no competing interest.

